# Placebo and Nocebo Responses in Multiple System Atrophy - a systematic review and meta-analysis of clinical trials

**DOI:** 10.1101/2024.11.05.24316747

**Authors:** Afonso Pessoa de Amorim, Filipe B Rodrigues, Ana Castro-Caldas, Joaquim J Ferreira

**Author notes:** **Corresponding author** Joaquim J Ferreira, Laboratório de Farmacologia Clínica e Terapêutica, Faculdade de Medicina de Lisboa, Av. Prof. Egas Moniz, 1649-028 Lisboa, Portugal.

## Abstract

**BACKGROUND:** Multiple System Atrophy (MSA) is a rapidly progressive and fatal neurodegenerative disease, with no effective treatment. Estimating the placebo and nocebo responses will help better design and interpret clinical trials.

*Objective:* To estimate the placebo and nocebo responses in MSA and explore their determinants.

**METHODS:** Electronic databases were searched up to November 2020. Randomized, blinded, placebo- or sham-controlled trials of patients with MSA were included if quantitative data were extractable on the placebo arm. The primary outcomes were: placebo response, defined as the within-group change from baseline, using any scale measuring motor outcomes; and nocebo response, defined as the proportion of patients experiencing adverse effects in the placebo arm. Random-effects meta-analyses were used to pool data. Several predetermined subgroup analyses and metaregressions were performed. PROSPERO registration number: CRD42021222915.

**RESULTS:** We included 21 randomized controlled trials (614 participants). Pooled placebo response was an increase in the Unified MSA Rating Scale (UMSARS) parts I and II of 9.09 points (95% CI 7.78 to 10.31, I^2^=94.00%, 9 studies, 304 participants). Pooled nocebo response was 63,88% (CI 95% 41.15 to 84.05, I^2^ =93.03%, 13 studies, 331 participants). Both placebo and nocebo responses were greater in trials with longer duration, whereas nocebo response was also higher in studies testing pharmacological interventions when compared with non-pharmacological interventions.

**CONCLUSIONS:** There may be a favorable response associated with the placebo, but this data needs to be compared with a “no treatment group” in order to validate its real impact. The nocebo response is high and should be considered in future clinical trial design and interpretation.

## Introduction

Multiple System Atrophy (MSA) is a progressive and fatal neurodegenerative disease secondary to oligodendroglial cytoplasmatic inclusions of misfolded a-synuclein protein.^1^ Its hallmark manifestations are autonomic dysfunction with different degrees and combinations of parkinsonism, pyramidal signs or cerebellar ataxia.^2^ There is an unmet need for efficacious symptomatic and disease-modifying interventions for people with MSA; however clinical trials thus far have revealed unsatisfactory results.

The placebo effect is described as the impact of the expectation of receiving a therapeutic intervention and this effect may hinder signal detection in clinical trials^3^. However, no studies systematically evaluate its magnitude and characteristics in MSA. Here we describe the placebo and nocebo responses in randomized placebo-controlled trials of patients with MSA with the aim of informing future trial design and interpretation.

## Methods

This report follows the PRISMA guidelines^4^ and the protocol was prospectively registered with PROSPERO (CRD42021222915). Full methodological details can be found in Supplementary Materials.

### Eligibility criteria

Randomized controlled trials (RCTs), of parallel or crossover design, double- or single-blinded, studying the effects of therapeutic interventions in people with clinical diagnosis of MSA; trials had to be published or registered with results; all studies had to be placebo or sham controlled; participants had to be adults (≥ 18 years of age). Due to small study bias, studies including less than five participants with MSA were excluded. There were no restrictions on disease state, number of centers, language, setting, duration, or year of publication. Studies had to report quantitative data on at least one of the following outcomes within the placebo arm:

#### Primary Efficacy Outcome

“placebo effect response”, defined as the within-group change from baseline, using any rating scale measuring the motor domain.

#### Primary Safety Outcome

“nocebo effect response”, defined as the proportion of patients experiencing adverse effects (AE) in the placebo arm.

#### Secondary Efficacy Outcomes

within-group change from baseline, using any rating scale measuring the following domains: functional ability, autonomic features, quality of life, and patient or clinical subjective impression of change.

#### Secondary Safety Outcomes

proportion of withdrawals and of patients experiencing serious adverse effects (SAE) in the placebo arm.

### Information Sources and Search

MEDLINE, Embaseand the Cochrane Register of Controlled Trials (CENTRAL) and NIH’s ClinicalTrials.gov from inception to November 2020. Search strategies are presented in Supplementary Materials.

### Selection Process

Two independent reviewers screened references and assessed full-texts for eligibility. Disagreements were resolved with discussion or by a third author.

### Data collection process

Two independent reviewers extracted data to a piloted electronic data extraction form that was cross-checked for accuracy.

### Risk of bias in individual studies

We used the Cochrane Risk of Bias tool^5^. The following items were rated low, unclear or high risk of bias : random sequence generation, allocation concealment, blinding of participants and personnel, blinding of outcome assessment, incomplete outcome data, and selective reporting. Selective reporting was classified as high risk of bias if the study did not report on all our predefined outcomes. The study overall was classified as moderate if at least one item was at moderate risk, and as high risk if any item was deemed as high risk. Two authors independently assessed each domain and disagreements were solved by discussion or by a third author.

### Statistical methods

Only placebo arm data were collected and analyzed. Placebo and nocebo data were derived from the last measured within-group difference or proportion in the placebo arm, respectively. Data was combined using random-effects meta-analyses techniques, namely the Dersimonian-Laird inverse-variance weighted model (Dersimonian and Laird 1986) for continuous and categorical variables. Permutational meta-regressions with 1,000 repetitions were used to explore association of the studied outcomes with continuous and categorical variables (i.e. meta-regression and subgroup analyses, respectively). Heterogeneity between results was assessed using the I^2^. Statistical package Stata 16.0 (Houston, Texas) was used and results are presented with a 95% confidence interval (95% CI).

## Results

The electronic search returned 952 records, 45 references were subject to full text review, and 21 studies^6–25^ were included. Full search results, characteristics of included and excluded studies and risk of bias can be found in the Supplementary Materials.

### Placebo responses

#### Primary outcome

Seventeen studies (574 patients) reported on MSA severity assessment scales, nine (304 patients) using Unified MSA Rating Scale (UMSARS) part I and II, and 8 (270 patients) using other tools. The overall pooled placebo response standardized mean difference was 1.04 (95% CI 0.59 to 1.49, n = 574, 17 studies; Supplementary Materials) and 9.09 UMSARS part I and II points (95% CI 7.78 to 10.31; 9 studies, n=304; Figure 1, Table 1) Subgroup analysis revealed a positive association with trial duration (Table 1). Statistical heterogeneity was not explained by the predefined subgroup and sensitivity analysis.

**Table 1.** Placebo (UMSARS part I and II) and nocebo (adverse events) responses subgroup and sensitivity analyses. 95% CI, 95% confidence interval.

|  | Nocebo Response<br>Proportion of Adverse Effects |  |  | Placebo Response<br>Motor Evaluation - UMSARS-I+II |  |  |
| --- | --- | --- | --- | --- | --- | --- |
|  | Percentage (95% CI) | p value | I2 (%) | Effect Size (95% CI) | p value | I2 (%) |
| **Global Results** |  |  |  |  |  |  |
|  | 63.88 (41.15 to 84.05) | p<0.001 | 93.03 % | 9.09 (7.78 to 10.31) | p<0.001 | 94.00 % |
| **Result Consistency** |  |  |  |  |  |  |
| **Publication Status** |  |  |  |  |  |  |
| Published | 92.96 (38.37 to 84.77) | p<0.001 | 93.61 % | 9.64 (8.36 to 10.91) | p<0.001 | 94.40 % |
| Unpublished | 73.68 (51.21 to 88.19) | p<0.001 | n/a | 4.70 (2.32 to 7.07) | p<0.001 | n/a |
| Difference | -12.27 (-97.50 to 72.96) | p=0.766 | 99.60 % | 5.62 (-6.33 to 17.58) | p=0.329 | 94.37 % |
| **Data Imputation** |  |  |  |  |  |  |
| Imputation | 70.70 (17.63 to 1.00) | p=0.001 | 0.00% | n/a | n/a | n/a |
| No Imputation | 61.81 (35.82 to 84.95) | p<0.001 | 94.06 % | n/a | n/a | n/a |
| Difference | 3.33 (-51.52 to 58.20) | p=0.918 | 99.48 % | n/a | n/a | n/a |
| **Study Design** |  |  |  |  |  |  |
| **Follow-Up (weeks)** |  |  |  |  |  |  |
| Coef | 1.40 (0.83 to 1.96) | p<0.001 | 86.46 % | 0.22 (0.01 to 0.42) | p=0.020 | 91.50 % |
| **Cross-over vs Parallel** |  |  |  |  |  |  |
| Cross-Over | 47.73 (26.11 to 69.71) | p<0.001 | 0.00% | n/a | n/a | n/a |
| Parallel | 67.45 (41.47 to 89.10) | p<0.001 | 94.61 % | n/a | n/a | n/a |
| Difference | -16.88 (-73.07 to 39.30) | p=0.520 | 99.61 % | n/a | n/a | n/a |
| **Intervention** |  |  |  |  |  |  |
| Pharmacological | 78.60 (64.72 to 90.10) | p<0.001 | 81.12 % | n/a | n/a | n/a |
| Non-Pharmacological | 00.00 (0.00 to 4.90) | p=1.000 | 0.00% | n/a | n/a | n/a |
| Difference | 77.63 (47.53 to 107.72) | p=0.015 | 87.60 % | n/a | n/a | n/a |
| **International** |  |  |  |  |  |  |
| International | 78.10 (69.69 to 85.55) | p<0.001 | 61.23 % | 11.71 (6.22 to 17.21) | p<0.001 | 92.70 % |
| Non-international | 59.50 (30.36 to 85.78) | p<0.001 | 93.87 % | 8.35 (6.58 to 10.13) | p<0.001 | 92.90 % |
| Difference | 23.62 (-36.43 to 83.66) | p=0.363 | 99.59 % | 3.09 (-5.36 to 11.54) | p=0.358 | 92.84 % |
| **Multi-center** |  |  |  |  |  |  |
| Multi-centric | 78.04 (63.19 to 90.21) | p<0.001 | 81.84 % | 9.47 (7.66 to 11.27) | p<0.001 | 94.70 % |
| Single-Centric | 37.44 (0.00 to 92.98) | p<0.001 | 95.45 % | 8.30 (7.92 to 8.68) | p<0.001 | n/a |
| Difference | 35.47 (-5.05 to 75.99) | p=0.130 | 99.24 % | 1.63 (-11.23 to 14.48) | p=1,000 | 94.74 % |
| **Year of Publication** |  |  |  |  |  |  |
| Coef | 0.04 (-2.66 to 2.73) | p=0.949 | 99.55 % | -0.45 (-1.35 to 0.45) | p=0.181 | 93.97 % |
| **Allocation Ratio** |  |  |  |  |  |  |
| Coef per % change | -0.01 (-0.03 to 0.02) | p=0.530 | 99.69 % | 0.00 (0.00 to 0.00) | p=0.236 | 94.67 % |
| **Population Characteristics** |  |  |  |  |  |  |
| **% males** |  |  |  |  |  |  |
| Coef per % change | 1.07 (-0.39 to 2.53) | p=0.213 | 99.51 % | 0.19 (-0.19 to 0.57) | p=0.412 | 94.52 % |
| **Mean Age (years)** |  |  |  |  |  |  |
| Coef | 2.52 (-1.35 to 6.39) | p=0.119 | 99.01 % | 0.51 (-0.94 to 1.96) | p=0.495 | 94.72 % |
| **Mean Motor Scale (units)** |  |  |  |  |  |  |
| Coef | 0.01 (-2.39 to 2.41) | p=0.940 | 91.76 % | -0.19 (-0.79 to 0.40) | p=0.727 | 91.14 % |

**Figure 1.**
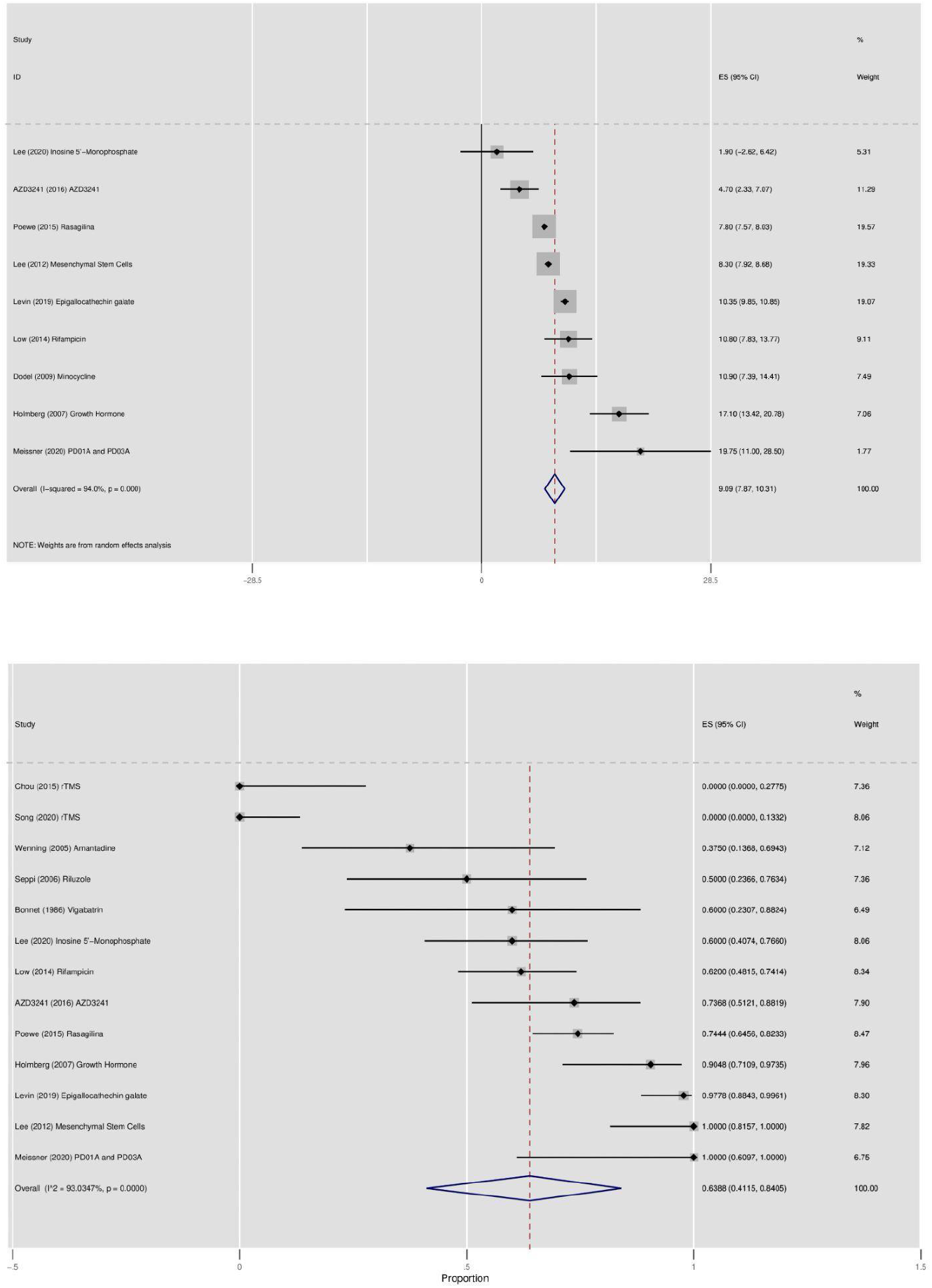
Forest plots for placebo (upper panel, UMSARS I & II) and nocebo (lower panel,) responses. ES, Effect size; 95% CI, 95% confidence interval.

#### Secondary outcomes

Four studies (250 participants) reported on Functional Ability, 3 (49 participants) reporting on the Unified Parkinson’s Disease Rating Scale (UPDRS) part II. The pooled placebo response was 2.52 UPDRS part II points (95% CI 0.45 to 4.61, n = 49, 3 studies; Supplementary Materials). Six studies (187 participants) reported on autonomic evaluations, however using different assessment techniques not possible to compare. Three studies (146 participants) used the Composite Autonomic Symptom Score (COMPASS), placebo response of 16.65 (95% CI 2.58 to 30.73; n = 146, 3 studies; Supplementary Materials). Not enough information was available to evaluate other domains. For the evaluated domains, planned subsequent analyses were not performed due to data sparsity.

### Nocebo responses

#### Primary outcome

13 studies (331 patients) reported the proportion of patients experiencing AE in the placebo arm. Overall, 63,88% (95% CI 41.15 to 84.05 n=331, 13 studies; Figure 1 and Table 1) of patients experienced AE. This proportion was higher when comparing pharmacological to non-pharmacological interventions and positively associated with trial duration (Table 1). Statistical heterogeneity was not explained by the predefined subgroup and sensitivity analysis.

#### Secondary outcomes

13 studies (366 participants) reported on withdrawals and 10 studies (291 participants) on SAE. 13,32% (95% CI 7.54 to 20.13, n=366, 13 studies; Supplementary Materials) of the trial participants withdrew study participation and 13,80% (95% CI 5.77 to 24.02, n=291, 10 studies; Supplementary Materials) reported SAE. Meta-regression and subgroup analyses can be found in the Supplementary Materials.

## Discussion

To our knowledge this work is the first systematic review evaluating the placebo and the nocebo responses in MSA. Our analyses showed a mean placebo response in MSA clinical trials of an increase (worsening) of 9.09 points in UMSARS part I and II. This effect is higher in studies with a longer duration. We also documented that 63.88% placebo-treated participants report AEs, with higher proportions in longer studies and in pharmacology studies.

This response in the placebo arm is expected, given the rapidly progressive nature of MSA. However, our results are limited by the available evidence, both in terms of the amount of eligible data and the absence of a “no treatment group”^26^. Using meta-regression we estimate an average annual increase of 11.44 points in UMSARS part I and II. Wenning et al. natural history cohort^27^ observed an annual worsening of 14.7 points, which among other explanations, could support the existence of a placebo response in our data (i.e., clinical trial participants on average progress lower than natural history study participants). If we assume this hypothesis to be true, the difference between the annualized rate of change in placebo arms and observational studies is larger than the estimated minimal clinical important decline for UMSARS part I and II of 3.0 points.^28^

The observed proportion of AE is aligned with previous studies and can also be confounded by the disease’s natural history. Expectedly, longer trials have a higher perceived dose exposure motivating larger nocebo responses.^29^ The differential effect noted in trials studying pharmacological interventions might be related with the follow-up duration as on average they are longer than non-pharmacological intervention trials.

There are limitations to our work. The reduced number of eligible studies limits our statistical power, weakening our ability to detect signals and the precision of the estimates. We refrained from including small studies and analyzing outcome domains with scarce data. Although our outcomes and analyses were predefined, we runned multiple analyses, increasing the risk of false positives. For these two reasons, secondary endpoints and subgroup or meta-regressions should be interpreted with caution. Cross-over studies were included, where a potential carry-over effect can overestimate the nocebo response. However we did not observe this effect on subgroup-analysis. Finally, the absence of data from prospectively and contemporaneous no-treatment-group arms precludes us from making assertive statements about placebo and nocebo effects, as these effects can be confounded by the Hawthorne effect (i.e. the effect of changing behaviors due to the awareness of being observed.^30^

We believe these observations could be of interest for planning and interpreting future clinical trials of people with MSA.

## Data Availability

All data produced in the present work are contained in the manuscript

## Bibliography

1. Stefanova N, Bücke P, Duerr S, Wenning GK. Multiple system atrophy: an update. 2009;8:7.

2. Wenning GK, Stankovic I, Vignatelli L, et al. The Movement Disorder Society Criteria for the Diagnosis of Multiple System Atrophy. Mov Disord. 2022;37(6):1131–1148. doi:10.1002/mds.29005

3. Požgain I, Požgain Z, Degme D. PLACEBO AND NOCEBO EFFECT: A MINI-REVIEW. Psychiatr Danub. 26(2):8.

4. Moher; D, Liberati A, Tetzlaff J, Altman D, PRISMA Group T. Preferred Reporting Items for Systematic Reviews and Meta-Analyses: The PRISMA Statement.

5. Higgins J, Altman D. Assessing Risk of Bias in Included Studies. In: Cochrane Handbook for Systematic Reviews of Interventions:. ; 2008:187–241.

6. Bensimon G, Ludolph A, Agid Y, Vidailhet M, Payan C, Leigh PN. Riluzole treatment, survival and diagnostic criteria in Parkinson plus disorders: The NNIPPS Study. Brain. 2009;132(1):156–171. doi:10.1093/brain/awn291

7. Bonnet AM, Esteguy M, Tell G, Schechter PJ, Hardenberg J, Agid Y. A Controlled Study of Oral Vigabatrin (?-Vinyl GABA) in Patients with Cerebellar Ataxia. Can J Neurol Sci J Can Sci Neurol. 1986;13(4):331–333. doi:10.1017/S0317167100036672

8. Bordet R, Benhadjali J, Destée A, Belabbas A, Libersa C. Octreotide Effects on Orthostatic Hypotension in Patients with Multiple System Atrophy: A Controlled Study of Acute Administration. Clin Neuropharmacol. 1995;18(1):83–89. doi:10.1097/00002826-199502000-00012

9. Chou Y hui, You H, Wang H, et al. Effect of Repetitive Transcranial Magnetic Stimulation on fMRI Resting-State Connectivity in Multiple System Atrophy. Brain Connect. 2015;5(7):451–459. doi:10.1089/brain.2014.0325

10. Dodel R, Spottke A, Gerhard A, et al. Minocycline 1-year therapy in multiple-system-atrophy: Effect on clinical symptoms and [^11^ C] (R) -PK11195 PET (MEMSA-trial): Efficacy of Minocycline for Patients with MSA-P. Mov Disord. 2010;25(1):97–107. doi:10.1002/mds.22732

11. França C, de Andrade DC, Silva V, et al. Effects of cerebellar transcranial magnetic stimulation on ataxias: A randomized trial. Parkinsonism Relat Disord. 2020;80:1–6. doi:10.1016/j.parkreldis.2020.09.001

12. Friess E, Kuempfel T, Modell S, et al. Paroxetine treatment improves motor symptoms in patients with multiple system atrophy. Parkinsonism Relat Disord. 2006;12(7):432–437. doi:10.1016/j.parkreldis.2006.04.002

13. Holmberg B, Johansson JO, Poewe W, et al. Safety and tolerability of growth hormone therapy in multiple system atrophy: A double-blind, placebo-controlled study. Mov Disord. 2007;22(8):1138–1144. doi:10.1002/mds.21501

14. Kaufmann H, Freeman R, Biaggioni I, et al. Droxidopa for neurogenic orthostatic hypotension: A randomized, placebo-controlled, phase 3 trial. Neurology. 2014;83(4):328–335. doi:10.1212/WNL.0000000000000615

15. Lee PH, Lee JE, Kim HS, et al. A randomized trial of mesenchymal stem cells in multiple system atrophy. Ann Neurol. 2012;72(1):32–40. doi:10.1002/ana.23612

16. Jung Lee J, Han Yoon J, Jin Kim S, et al. Inosine 5’-Monophosphate to Raise Serum Uric Acid Level in Multiple System Atrophy (IMPROVE-MSA study). Clin Pharmacol Ther. Published online November 30, 2020:cpt.2082. doi:10.1002/cpt.2082

17. Levin J, Maaß S, Schuberth M, et al. Safety and efficacy of epigallocatechin gallate in multiple system atrophy (PROMESA): a randomised, double-blind, placebo-controlled trial. Lancet Neurol. 2019;18(8):724–735. doi:10.1016/S1474-4422(19)30141-3

18. Low PA, Robertson D, Gilman S, et al. Efficacy and safety of rifampicin for multiple system atrophy: a randomised, double-blind, placebo-controlled trial. Lancet Neurol. 2014;13(3):268–275. doi:10.1016/S1474-4422(13)70301-6

19. Meissner WG, Traon AP, Foubert-Samier A, et al. A Phase 1 Randomized Trial of Specific Active α-Synuclein Immunotherapies PD01A and PD03A in Multiple System Atrophy. Mov Disord. 2020;35(11):1957–1965. doi:10.1002/mds.28218

20. Saccà F, Marsili A, Quarantelli M, et al. A randomized clinical trial of lithium in multiple system atrophy. J Neurol. 2013;260(2):458–461. doi:10.1007/s00415-012-6655-7

21. Seppi K, Peralta C, Diem-Zangerl A, et al. Placebo-controlled trial of riluzole in multiple system atrophy. Eur J Neurol. 2006;13(10):1146–1148. doi:10.1111/j.1468-1331.2006.01452.x

22. Song P, Li S, Wang S, Wei H, Lin H, Wang Y. Repetitive transcranial magnetic stimulation of the cerebellum improves ataxia and cerebello-fronto plasticity in multiple system atrophy: a randomized, double-blind, sham-controlled and TMS-EEG study. Aging. 2020;12(20):20611–20622. doi:10.18632/aging.103946

23. Wenning GK. Placebo-Controlled Trial of Amantadine in Multiple-System Atrophy: Clin Neuropharmacol. 2005;28(5):225–227. doi:10.1097/01.wnf.0000183240.47960.f0

24. Wessel K, Hermsdorfer J, Deger K, et al. Double-blind Crossover Study With Levorotatory Form of Hydroxytryptophan in Patients With Degenerative Cerebellar Diseases. Arch Neurol. 1995;52(5):451–455. doi:10.1001/archneur.1995.00540290037015

25. AstraZeneca. A 12-Week, Multicenter, Randomized, Parallel-Group Study to Assess the Safety, Tolerability, Pharmacokinetics, Biomarker Effects, Efficacy, and Effect on Microglia Activation, as Measured by Positron Emission Tomography, of AZD3241 in Subjects With Multiple System Atrophy. clinicaltrials.gov; 2017. Accessed February 11, 2021. https://clinicaltrials.gov/ct2/show/NCT02388295

26. Ferreira JJ, Trenkwalder C, Mestre TA. Placebo and nocebo responses in other movement disorders besides Parkinson’s disease: How much do we know?: Knowledge of Placebo and Nocebo Responses Outside PD. Mov Disord. 2018;33(8):1228–1235. doi:10.1002/mds.113

27. Wenning GK, Geser F, Krismer F, et al. The natural history of multiple system atrophy: a prospective European cohort study. Lancet Neurol. 2013;12(3):264–274. doi:10.1016/S1474-4422(12)70327-7

28. Krismer F, Seppi K, Wenning GK, Abler V, Papapetropoulos S, Poewe W. Minimally clinically important decline in the parkinsonian variant of multiple system atrophy. Mov Disord. 2016;31(10):5.

29. Webster RK, Weinman J, Rubin GJ. A systematic review of factors that contribute to nocebo effects. Health Psychol. 2016;35(12):1334–1355. doi:10.1037/hea0000416

30. Rodrigues FB, Ferreira JJ. Strategies to minimize placebo effects in research investigations. In: International Review of Neurobiology. Vol 153. Elsevier; 2020:49–70. doi:10.1016/bs.irn.2020.04.002

