## Supplementary Data for "Placebo and Nocebo Responses in Multiple System Atrophy - a systematic review and meta-analysis of clinical trials"

**Title**

**Corresponding author**

### Abstract

***BACKGROUND:*** Multiple System Atrophy (MSA) is a rapidly progressive and fatal neurodegenerative disease, with no effective treatment. Estimating the placebo and nocebo responses will help better design and interpret clinical trials.

***Objective:*** To estimate the placebo and nocebo responses in MSA and explore their determinants.

***METHODS:*** Electronic databases were searched up to November 2020. Randomized, blinded, placebo- or sham-controlled trials of patients with MSA were included if quantitative data were extractable on the placebo arm. The primary outcomes were: placebo response, defined as the within-group change from baseline, using any scale measuring motor outcomes; and nocebo response, defined as the proportion of patients experiencing adverse effects in the placebo arm. Random-effects meta-analyses were used to pool data. Several predetermined subgroup analyses and metaregressions were performed. PROSPERO registration number: CRD42021222915.

***RESULTS:*** We included 21 randomized controlled trials (614 participants). Pooled placebo response was an increase in the Unified MSA Rating Scale (UMSARS) parts I and II of 9.09 points (95% CI 7.78 to 10.31, $I^2$=94.00%, 9 studies, 304 participants). Pooled nocebo response was 63,88% (CI 95% 41.15 to 84.05, $I^2$ =93.03%, 13 studies, 331 participants). Both placebo and nocebo responses were greater in trials with longer duration, whereas nocebo response was also higher in studies testing pharmacological interventions when compared with non-pharmacological interventions.

***CONCLUSIONS:*** There may be a favorable response associated with the placebo, but this data needs to be compared with a "no treatment group" in order to validate its real impact. The nocebo response is high and should be considered in future

clinical trial design and interpretation.

### Supplementary Materials

#### Full Methodology

**Standard protocol approvals and registrations:** We report this systematic review and meta-analysis according to the Preferred Reporting Items for Systematic Reviews and Meta-Analyses Guidelines.[1] The protocol was prospectively registered with PROSPERO (registration: CRD42021222915).

**Eligibility criteria:** We included randomized controlled trials (RCTs), of parallel or crossover design, double- or single-blinded, studying the effects of therapeutic interventions in people with MSA. We accepted trials that were published or registered with results. All studies had to be placebo or sham controlled. Participants had to be adults (i.e. ≥ 18 years of age) with a clinical diagnosis made by any physician, specialist or otherwise, of MSA. Due to small study bias, studies including less than five participants with MSA were excluded. There were no restrictions regarding the disease state, the number of recruitment centres, language, setting, duration, or year of publication. Studies had to report quantitative data on at least one of the following outcomes, measured by validated instruments, within the placebo arm:

***Primary Efficacy Outcome***: The primary efficacy outcome was "placebo effect response", defined as the within-group change from baseline, using any rating scale measuring the motor domain

***Primary Safety Outcome***: The primary safety outcome was "nocebo effect response", defined as the proportion of patients experiencing adverse effects in the placebo arm.

***Secondary Efficacy Outcomes***: The secondary safety outcomes were within-group change from baseline, using any rating scale measuring the following domains: functional ability, autonomic features, quality of life, and subjective impression of change rated by the patient or clinician.

***Secondary Safety Outcomes***: The secondary safety outcomes were proportion of withdrawals, and proportion of patients experiencing serious adverse effects in the placebo arm.

**Information Sources:** We searched MEDLINE, EMBASE and the Cochrane Register of Controlled Trials (CENTRAL) and FDA's Clinical Trials Database from inception to

November 2020. Searches through reference lists of located trials and of review articles were also conducted to increase the sensitivity of our research strategy.

**Search:** For the identification of studies considered for inclusion, a detailed search strategy was developed for each database searched. All search strategies included Multiple System Atrophy, and database-specific search filters were included to select RCT.[2–4] Below you will find the MEDLINE, CENTRAL and EMBASE strategy.

**Selection Process**: Two independent reviewers screened the references identified by the search strategy, reading each of the titles and abstracts and excluding references not applicable according to the eligibility criteria. If the abstract was not available for screening, we opted to retrieve the full text of such study. When disagreements arose, they were resolved through discussion or, when necessary, by a third author.Two independent reviewers assessed the full-text articles to see if the studies fulfilled the inclusion criteria. Again, disagreements were resolved through discussion or, if necessary, through the consultation of a third author.

**Data collection process**: Two independent reviewers extracted data to a predetermined and piloted electronic data extraction form. The forms were cross-checked for accuracy.

**Risk of bias in individual studies**: We used the Cochrane Risk of Bias tool to assess the overall methodological quality of each study[5]. This tool was constructed to incorporate several sources of bias such as selection bias, performance bias, detection bias, attrition bias, reporting bias and other biases. Effectively, we rated a low, unclear or high risk of bias to the seven following items: random sequence generation, allocation concealment, blinding of participants and personnel, blinding of outcome assessment, incomplete outcome data, and selective reporting; to what selective reporting regards, we classified as high risk of bias any study that did not report on all our predefined outcomes, both primary and secondary. As for general risk of bias, we classified as moderate risk a study that did not have all items classified as low risk, and as high risk a study that any item deemed as high risk. Two authors independently assessed each risk of bias domain and disagreements were solved by discussion and, if necessary, by a third author.

**Summary Measures:** For the study of placebo and nocebo responses, we only considered RCTs where the control arm was a placebo intervention. Furthermore, only the data regarding the placebo arm was collected and analysed. The placebo and nocebo data was derived from the last measured within-group response in the placebo arm. The effect measures chosen for continuous variables were the raw values or the effect size according to the Hedges g corrected for small-sample bias[6,7], and for dichotomous variables we used the relative frequencies in the form of percentages.

In case of insufficient reported data, we contacted authors as a first approach. In the lack of a positive response, absent mean changes from baseline and SDs were extracted from unpublished material. In some trials, SD was obtained from standard error or confidence intervals (CI). When such values were not reported, imputation methods were applied. If variances or standard errors or standard deviations of mean differences were not available, they were imputed using a change from baseline standard deviation using a correlation coefficient[8,9]. When only a baseline or a final visit measure of dispersion was available but not the corresponding final visit or baseline value, correspondingly, we imputed the missing value as equal to the existing one, and used a conservative correlation coefficient. When necessary, medians and ranges were converted to means and standard deviations according to the methods developed by Hozo et colleagues[10].For cross-over trials, we included data from the placebo period only. If placebo data was provided by treatment period we only included the first placebo period. If neither of these were available, we excluded the studies from the analyses.

We calculated the allocation ratio for each study dividing the number of patients allocated to placebo by the number of patients allocated to the

Data was combined using random-effects meta-analysis techniques, namely the Dersimonian-Laird (Dersimonian and Laird 1986) model for (inverse-variance) weighted analysis for continuous and categorical variables. Permutational meta-regressions with 1,000 repetitions were used to explore association with continuous and categorical variables (i.e. meta-regression and subgroup analyses).

Heterogeneity between results was assessed using the $I^2$ performed to quantify inconsistency across studies.

We performed the statistical analysis with the statistical package Stata 16.0 (Houston, Texas). All results are presented with a 95% confidence interval (95% CI).

**Additional analysis**: We performed subgroup analyses for the following areas, independently of the presence or not of significant heterogeneity: publication status; data imputation status; risk of bias; and study design (i.e. parallel versus cross-over). Meta-regressions were planned according to: year of publication, follow-up duration, allocation ratio, participants' stage and age, and other baseline characteristics. These analyses were exploratory.

### Search Strategies

#### MEDLINE

1 exp Multiple System Atrophy/ (3598)

2 MSA.ti,ab.

3 (multiple adj3 system$ adj3 atroph$).ti,ab.

4 (multisystem$ adj3 atroph$).ti,ab.

5 (striatonigral adj degeneration).ti,ab.

6 (shy-drager adj3 syndrome$).ti,ab.

7 (olivopontocerebellar adj3 atroph$).ti,ab.

8 (olivoponto adj3 cerebellar adj3 atroph$).ti,ab.

9 or/1-8

10 randomized controlled trial.pt.

11 controlled clinical trial.pt.

12 randomized.ab.

13 placebo.ab.

14 clinical trials as topic.sh.

15 randomly.ab.

16 trial.ti.

17 or/10-16

18 exp animals/ not humans.sh.

19 17 not 18

20 and/9,19

#### Embase

1 exp Shy Drager syndrome/

2 MSA.ti,ab.

3 (multiple adj3 system$ adj3 atroph$).ti,ab.

4 (multisystem$ adj3 atroph$).ti,ab.

5 (striatonigral adj degeneration).ti,ab.

6 (shy-drager adj3 syndrome$).ti,ab.

7 (olivopontocerebellar adj3 atroph$).ti,ab.

8 (olivoponto adj3 cerebellar adj3 atroph$).ti,ab.

9 or/1-8

10 (random$ or placebo$ or single blind$ or double blind$ or triple blind$).ti,ab.

11 RETRACTED ARTICLE/

12 or/10-11

13 (animal$ not human$).sh,hw.

14 (book or conference paper or editorial or letter or review).pt. not exp randomized controlled trial/

15 (random sampl$ or random digit$ or random effect$ or random survey or random regression).ti,ab. not exp randomized controlled trial/

16 or/13-15

17 12 not 16

18 and/9,17

#### CENTRAL

#1 MeSH descriptor: [Multiple System Atrophy] explode all trees

#2 multiple NEAR/3 system* NEAR/3 atroph*

#3 multisystem* NEAR/3 atroph*

#4 striatonigral NEAR/3 degeneration

#5 shy-drager NEAR/3 syndrome*

#6 olivopontocerebellar NEAR/3 atroph*

#7 olivoponto NEAR/3 cerebellar NEAR/3 atroph*

#8 #1 OR #2 OR #3 OR #4 OR #5 OR #6 OR #7 in Trials

### Full Search Results

**Publications:** The electronic search returned 952 records (274 from CENTRAL, 301 from MEDLINE, 377 from EMBASE, and none from other database sources), resulting in 621 records after all duplicates were removed. Through abstract screening, 565 records were removed, and after the application of inclusion and exclusion criteria to full-texts, 21 publications were included. Figure 1 shows the flowchart of the search. The detailed reason for exclusion for the 24 studies excluded in full-text review can be found below.

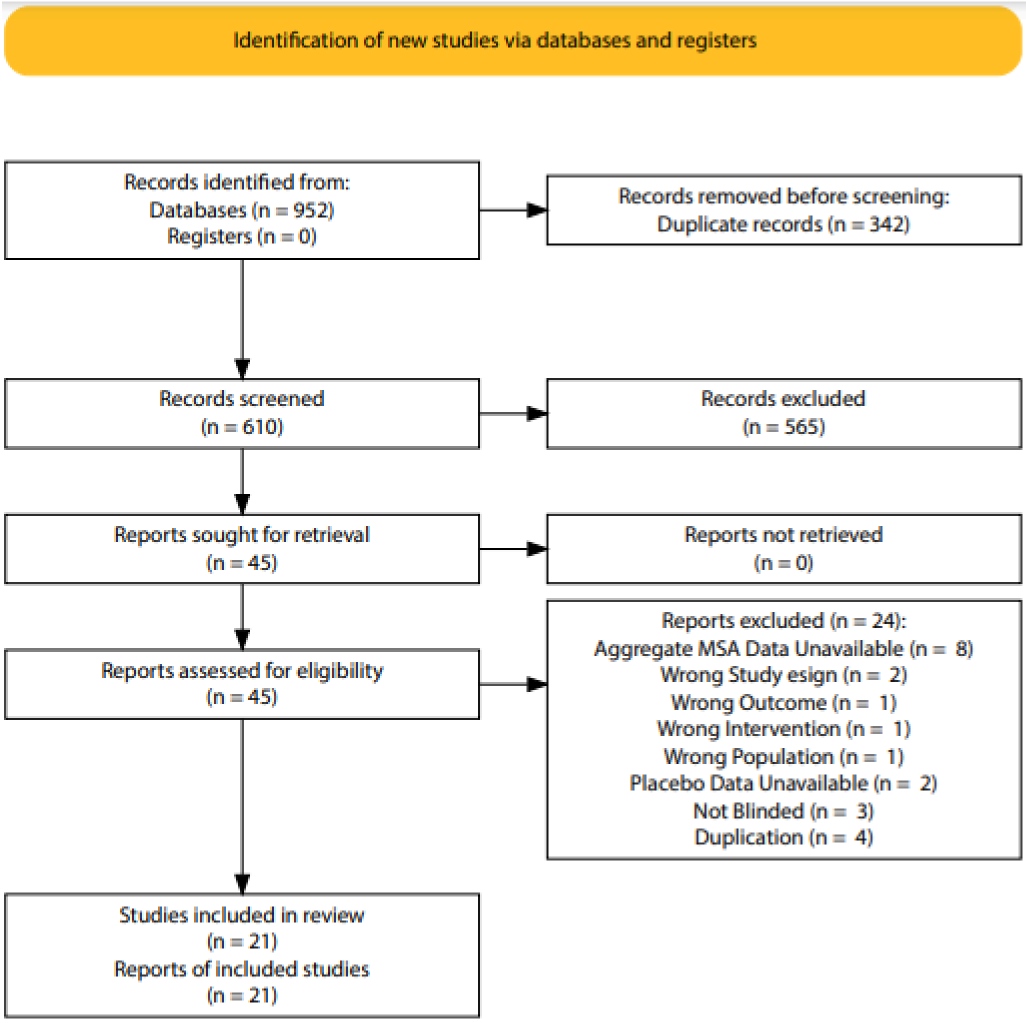


**Suppl Fig. 1** - Search flowchart.[11]

#### Trial Characteristics

**Publications**: A total of 21 studies, 20 publications[12–31] and 1 clinical trial registration with results[32], were included, with 16 studies[14–16,18,19,21–27,29,31–33] including only MSA patients and 5[12,13,17,20,30] including MSA and other conditions. All reports were written in English. A resume table with the study characteristics can be found in Appendix e-4.

**Study Design:** These trials were conducted between 1986 and 2020. Only 12 studies were multicentric[12,16,19,20,22–25,29–32], and of these only 5 were international[12,16,19,20,31], and the remaining 9 were single-center[13–15,17,18,21,26,27,33]. All the 21 trials were controlled with placebo[12–14,16,18–27,29–32] (18 studies, n=571) or sham[15,17,33] (3 studies, n=43) intervention; 6 trials were cross-over[13,14,17,27,29,30] (33,3%) and the remaining[12,15,16,18–26,31–33] (n=15, 66,7%) had a parallel design. Regarding outcomes, 13 studies (61,9%)[15–19,21,23,24,27,29–31,33] had a primary motor outcome, 5 (23,8%)[12,22,25,26,32] had safety and tolerability as their primary outcome and 2 (9,5%)[14,20] had autonomic features as their primary outcome; 1 study (4,8%)[13] had cerebellar symptomatology as its primary outcome.

**Study Participants**: The included studies enrolled a total of 1,221 participants, with 614 (50,2%) in the placebo arm. Regarding the participants included in the placebo arm, among those studies which declared such information (19 studies, n=596), 60% were males (n=359). The mean age in the placebo arms varied between 44 years[13] and 71 years[14]. Among the studies which provided a mean baseline evaluation of motor symptoms in total UMSARS score (I + II), the variation was between 27[24] and 55[26].

**Therapeutic Interventions**: The majority of clinical trials studied pharmacological interventions[12–14,16,18–27,29–32] (N=18, 85,7%) whereas only 3 trials[15,17,33] (14,3%) assessed non-pharmacological studies. (Table 1).

| Intervention | Number of Studies (%) |
|---|---|
| Amantidine[29] | 1 |
| AZD3241[32] | 1 |
| Droxidopa[20] | 1 |
| Epigallocathechin gallate[23] | 1 |
| Growth-Hormone[19] | 1 |
| Hydroxytryptophan[30] | 1 |
| Inosine 5'-Monophosphate[22] | 1 |
| Lithium[26] | 1 |

| | |
|---|---|
| Mesenchymal Stem Cells[21] | 1 |
| Minocycline[16] | 1 |
| Octreotide[14] | 1 |
| Paroxetin[18] | 1 |
| PD01A/PD03A[25] | 1 |
| Rasagiline[31] | 1 |
| Rifampicin[24] | 1 |
| Riluzole[12,27] | 2 |
| Repetitive transcranial magnetic stimulation[15,17,28] | 3 |
| Vigabatrin[13] | 1 |

Suppl Table 1 - Interventions included in the included studies

**Risk of Bias:** The extent of the methodological detail reported in the trials was not consistent, and several quality indications were not entirely considered in many publications (fig. 1). Only 12 studies[12,16,17,19–26,31] (57,1%) clearly described the process of random sequence generation; no studies were deemed to be at high risk, and 9 studies[13–15,18,27,29,30,32,33] (42,9%) were considered to be at an unclear risk for this criteria.

Regarding the allocation concealment process, only 11 studies[12,16,17,19–24,26,31] (52,4%) described an adequate process and therefore were rated as having a low risk of bias, whereas the remaining 10 studies[13–15,18,25,27,29,30,32,33] were assessed as having an unclear risk (47,6%).

As for blinding of participants, 20 studies[12–26,29,31–33] (95,2%) were deemed as having a low risk of bias; the other study[27] (4,8%) failed to provide an adequate blinding, being therefore considered at a high risk of bias; concerning the blinding of outcome assessment, all 21 studies (100%) were considered at a low risk of bias.

Incomplete assessment of outcomes was evaluated to be at a low risk on only 9 trials[14,15,17,20,27,29,30,32,33] (42,8%), whereas the remaining had an unclear[21,24] (n=2, 9.5%) or high[12,13,16,18,19,22,23,25,26,31] (n=10, 47.6%) risk of bias.

Finally, regarding selective outcome reporting data, all the 21 studies (100%) were deemed at a high risk of bias, because none reported all the criteria that we deemed necessary to our evaluation.

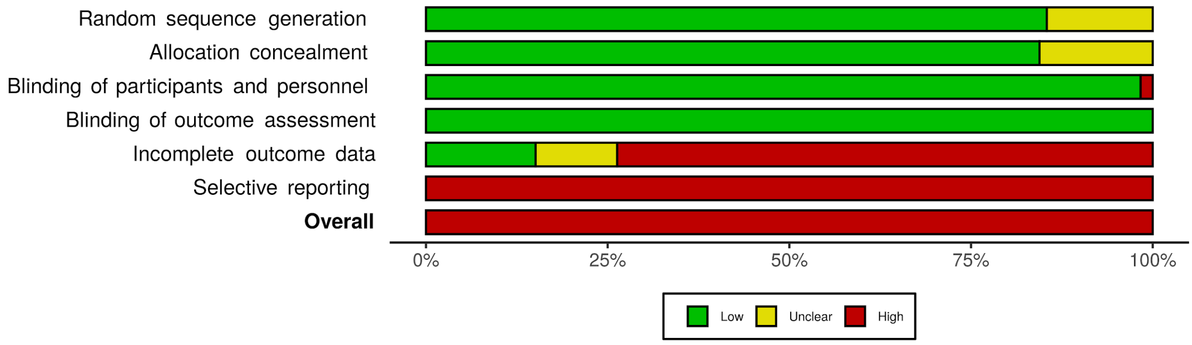


**Suppl Fig. 2.** Review author's judgements about each bias, presented as percentages across all studies.[34]

### Included Trials Characteristics

| Article | Intervention | Trial Design | n total | n placebo | Follow-up | % males | Mean age | Motor scale | Mean motor classification |
|---|---|---|---|---|---|---|---|---|---|
| Bensimon 2008[12] | Riluzole | Parallel, double-blinded, multicentric and international | 404 | 201 | 156 | 54,7% | 62,26 | Short Motor Disability Scale | 6,1 |
| Bonnet 1986[13] | Vigabatrin | Cross-over, double-blinded and single centric | 5 | 5 | 16 | 40% | 44 | . | . |
| Bordet 1995[14] | Ocreotide | Cross-over, double-blinded and single centric | 9 | 9 | immediate | 33,3% | 71 | . | . |
| Chou 2015[15] | Repetitive Transcranial Magnetic Stimulation | Parallel, double-blinded and single centric | 19 | 10 | 2 | 52,6% | 55 | UMSARS-II | 20 |
| Dodel 2009[16] | Minocycline | Parallel, double-blinded, multicentric and international | 63 | 31 | 48 | 46% | 62,02 | UMSARS-I+II | 42,99 |
| França 2020[17] | Repetitive Transcranial Magnetic Stimulation | Cross-over, double-blinded and single centric | 8 | 8 | 8 | 37,5% | 61,1 | Scale for the Assessment and Rating of Ataxia | 14 |
| Friess 2006[18] | Paroxetine | Parallel, double-blinded and single centric | 20 | 11 | 2 | 50% | 65,05 | UPDRS | 27,78 |
| Holmberg 2007[19] | Growth Hormone | Parallel, double-blinded, multicentric and international | 33 | 21 | 52 | 60,5% | 60,1 | UMSARS-I+II | 49,9 |
| Kaufmamn 2014[20] | Droxidopa | Parallel, double-blinded, multicentric and international | 33 | 11 | 1 | . | . | . | . |
| Lee 2012[21] | Mesenchymal Stem Cells | Parallel, double-blinded and single centric | 33 | 17 | 51 | 63,6% | 54,95 | UMSARS-I+II | 39,79 |
| Lee 2020[22] | Inosine | Parallel, double-blinded and | 55 | 25 | 24 | 54,5% | 63,1 | UMSARS-I+II | 53,28 |

| | | | | | | | | | |
|---|---|---|---|---|---|---|---|---|---|
| | 5'-Monophosphate | multicentric | | | | | | | |
| Levin 2019[23] | Epigallocathechin galate | Parallel, double-blinded and multicentric | 92 | 45 | 52 | 59,8% | 63,1 | UMSARS-I+II | 43,01 |
| Low 2014[24] | Rifampicin | Parallel, double-blinded and multicentric | 100 | 50 | 52 | 61% | 61 | UMSARS-I+II | 28,5 |
| Meissner 2020[25] | PD01A and PD03A | Parallel, double-blinded and multicentric | 30 | 6 | 52 | 63,3% | 59, | UMSARS-I+II | 59,5 |
| Poewe 2015[31] | Rasagiline | Parallel, double-blinded, multicentric and international | 174 | 90 | 48 | 57,5% | 65 | UMSARS-I+II | 37,22 |
| Saccá 2013[26] | Lithium | Parallel, double-blinded and single centric | 9 | 5 | 52 | 77,8% | 64 | UMSARS-I+II | 55 |
| Seppi 2006[27] | Riluzole | Cross-over, double-blinded and single centric | 10 | 10 | 12 | 30% | 62 | UPDRS-III | 38 |
| Song 2020[33] | Repetitive Transcranial Magnetic Stimulation | Parallel, double-blinded and single centric | 50 | 25 | 1 | 58% | 53,15 | Scale for the Assessment and Rating of Ataxia | 18,35 |
| Wenning 2005[29] | Amantadine | Cross-over, double-blinded and multicentric | 8 | 8 | 7 | 75% | 64,7 | UPDRS-III | 35 |
| Wessel 1995[30] | Hydroxytryptophan | Cross-over, double-blinded and multicentric | 7 | 7 | 90 | . | . | . | .. |
| AZD3241 2016[32] | ADZ3241 | Parallel, double-blinded and multicentric | 59 | 19 | 12 | 70,7% | 59 | . | . |

### Trials excluded in Full Text Review and reason

| Article | Intervention | Reason for exclusion |
|---|---|---|
| Armstrong 1991[35] | Ocreotide | No information regarding blinding of patients, therefore no assurance that there was any. |
| Aschoff 1996[36] | Physostigmine | Absence of isolated data for the MSA group |
| Bier 2003[37] | Ondaserton | Absence of isolated data for the MSA group |
| Botez 1996[38] | Amantadine Hydrochloride | Wrong outcome - only measuring reaction and movement time, without using any comparable scales. |
| Chaudhuri 1994[39] | Alcohol | Study had the intent of measuring the effect of alcohol ingestion, rather than studying any intervention therapy |
| Chou 2015[40] | Repetitive Transcranial Magnetic Ressonance | Duplicate from another study already included.[15] |
| Fouad-Tarazi 1995 [41] | Midodrine | The study only included 1 patient with MSA, therefore not meeting the minimum number of MSA subjects |
| Freeman 1996[42] | DL-DOPS | Absence of isolated data for the MSA group |
| Freeman 1999[43] | DL-DOPS | Absence of isolated data for the MSA group |
| Hussain 2001[44] | Sildenafil | Absence of isolated data for the MSA group |
| Isaacson 2016[45] | Droxidopa | Absence of isolated data for the MSA group |
| Jordan 2002[46] | COMT | No information regarding blinding of patients, therefore no assurance that there was any. |
| Jordan 1998[47] | Multiple vasopressor agents | Absence of data for the placebo group. |
| Kaufman 2003[48] | L-DOPS | Absence of isolated data for the MSA group |

| | | |
|---|---|---|
| Lee 2008[49] | Autologous Mesenchymal Stem Cells | Absence of any sort of blinding. |
| Lipp 2003[50] | Riluzole | Absence of any sort of blinding. |
| Mancini 2003[51] | Botulinum Toxin Type A | Absence of isolated data for the MSA group. |
| Mullen 2018[52] | AZD3241 | Only abstract available; information duplicated from another study, already included.[32] |
| Park 2016[53] | Mesenchymal Stem Cells | Duplicate from another study already included.[21] |
| Pollak 1987[54] | Apomorphine and Domperidone | The study only included 2 patients with MSA, therefore not meeting the minimum number of MSA subjects |
| Singer 2014[55] | Rifampicin | Duplicate from another study already included.[24] |
| Singer 2006[56] | Pyridostigmine | Absence of isolated data for the MSA group. |
| Sunwoo 2014[57] | Mesenchymal Stem Cells | Longitudinal non-blinded study. |
| Wang 2016[58] | Repetitive Transcranaial Magnetic Stimulation | Duplicate from another study already included.[40] |

### Efficacy Outcomes

#### Subgroup analysis for Primary Efficacy Outcomes

| | Placebo Response Motor Evaluation - UMSARS-I+II | | | Placebo Response Motor Evaluation - smd | | |
|---|---|---|---|---|---|---|
| | Effect Size (95% | p value | I2 (%) | Effect Size (95% | p value | I2 (%) |
| **Global Results** | | | | | | |
| | 9.09 (7.78 to 10. | p<0.001 | 94.00% | 1.04 (0.59 to 1.4 | p<0.001 | 97.3% |
| **Result Consistency** | | | | | | |
| **Publication Status** | | | | | | |
| Published | 9.64 (8.36 to 10. | p<0.001 | 94.40% | 1.08 (0.60 to 1.5 | p<0.001 | 97.40% |
| Unpublished | 4.70 (2.32 to 7.0 | p<0.001 | n/a | 0.58 (0.26 to 0.9 | p<0.001 | n/a |
| Difference | 5.62 (-6.33 to 17 | p=0.329 | 94.37% | 0.60 (-3.85 to 5.0 | p=0.714 | 97.44% |
| **Data Imputation** | | | | | | |
| Imputation | n/a | n/a | n/a | n/a | n/a | n/a |
| No Imputation | n/a | n/a | n/a | n/a | n/a | n/a |
| Difference | n/a | n/a | n/a | n/a | n/a | n/a |
| **Study Design** | | | | | | |
| **Follow-Up (weeks)** | | | | | | |
| Coef | 0.22 (0.01 to 0.4 | p= 0.020 | 91,50% | 0.01 (-0.02 to 0.3 | p=0.449 | 97.40% |
| **Cross-over vs Parallel** | | | | | | |
| Cross-Over | n/a | n/a | n/a | 00 (-0.21 to 0.20) | p=0.968 | 0.00% |
| Parallel | n/a | n/a | n/a | 87 (0.84 to 1.93) | p<0.001 | 97.80% |
| Difference | n/a | n/a | n/a | -1.51 (-3.84 to 0. | p=0.135 | 97.28% |
| **Intervention** | | | | | | |
| Pharmacological | n/a | n/a | n/a | 32 (0.81 to 1.84) | p<0.001 | 97.5% |
| Non-Pharmacological | n/a | n/a | n/a | 15 (-0.39 to 0.09) | p=0.224 | 28.4% |
| Difference | n/a | n/a | n/a | 1.60 (-1.00 to 4.2 | p=0.067 | 97.14% |
| **International** | | | | | | |
| International | 1 (6.22 to 17.21) | p<0.001 | 92.70% | .84 (0.57 to 3.11) | p=0.005 | 98.9% |
| Non-international | 8.35 (6.58 to 10. | p<0.001 | 92.90% | 78 (0.29 to 1.26) | p=0.002 | 95.6% |
| Difference | 3.09 (-5.36 to 11. | p=0.358 | 92.84% | 0.92 (-1.50 to 3.3 | p=0.325 | 97.26% |
| **Multi-center** | | | | | | |
| Multi-centric | 47 (7.66 to 11.27 | p<0.001 | 94.70% | 23 (0.67 to 1.80) | p<0.001 | 97.7% |
| Single-Centric | 8.30 (7.92 to 8.6 | p<0.001 | n/a | 61 (-0.08 to 1.29) | p=0.083 | 93.9% |
| Difference | 1.63 (-11.23 to 1 | p=1,000 | 94.74% | 0.31 (-1.90 to 2.5 | p=0.536 | 97.13% |
| **Year of Publication** | | | | | | |
| Coef | -0.45 (-1.35 to 0. | p=0.181 | 93.97% | 0.05 (-0.10 to 0.2 | p=0.465 | 97.43% |
| **Allocation Ratio** | | | | | | |
| Coef per % change | 0.00 (0.00 to 0.0 | p=0.236 | 94.67% | 0.00 (0.00 to 0.0 | p=0.752 | 98.00% |
| **Population Characteristics** | | | | | | |
| **% males** | | | | | | |
| Coef per % change | 0.19 (-0.19 to 0.5 | p=0.412 | 94.52% | 0.03 (-0.05 to 0.1 | 0=0.401 | 97.52% |
| **Mean Age (years)** | | | | | | |
| Coef | 0.51 (-0.94 to 1.9 | p=0.495 | 94.72% | 0.03 (-0.27 to 0.3 | p=0.575 | 97.29% |
| **Mean Motor Scale (units)** | | | | | | |
| Coef | -0.19 (-0.79 to 0. | p=0.727 | 91.14% | -0.05 (-0.34 to 0. | p=0.690 | 98.61% |

#### Forest Plot for Autonomic Function

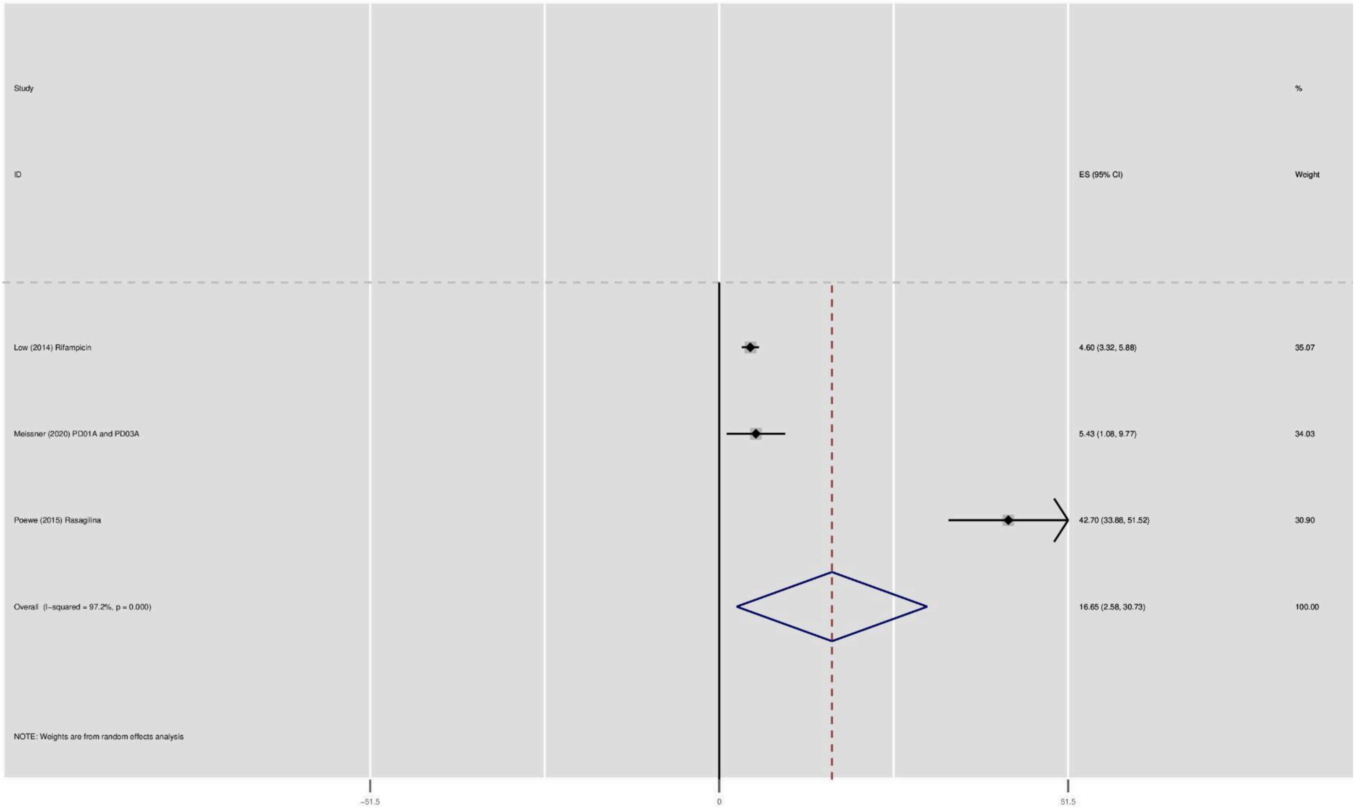


#### Forest Plot for Functional Ability

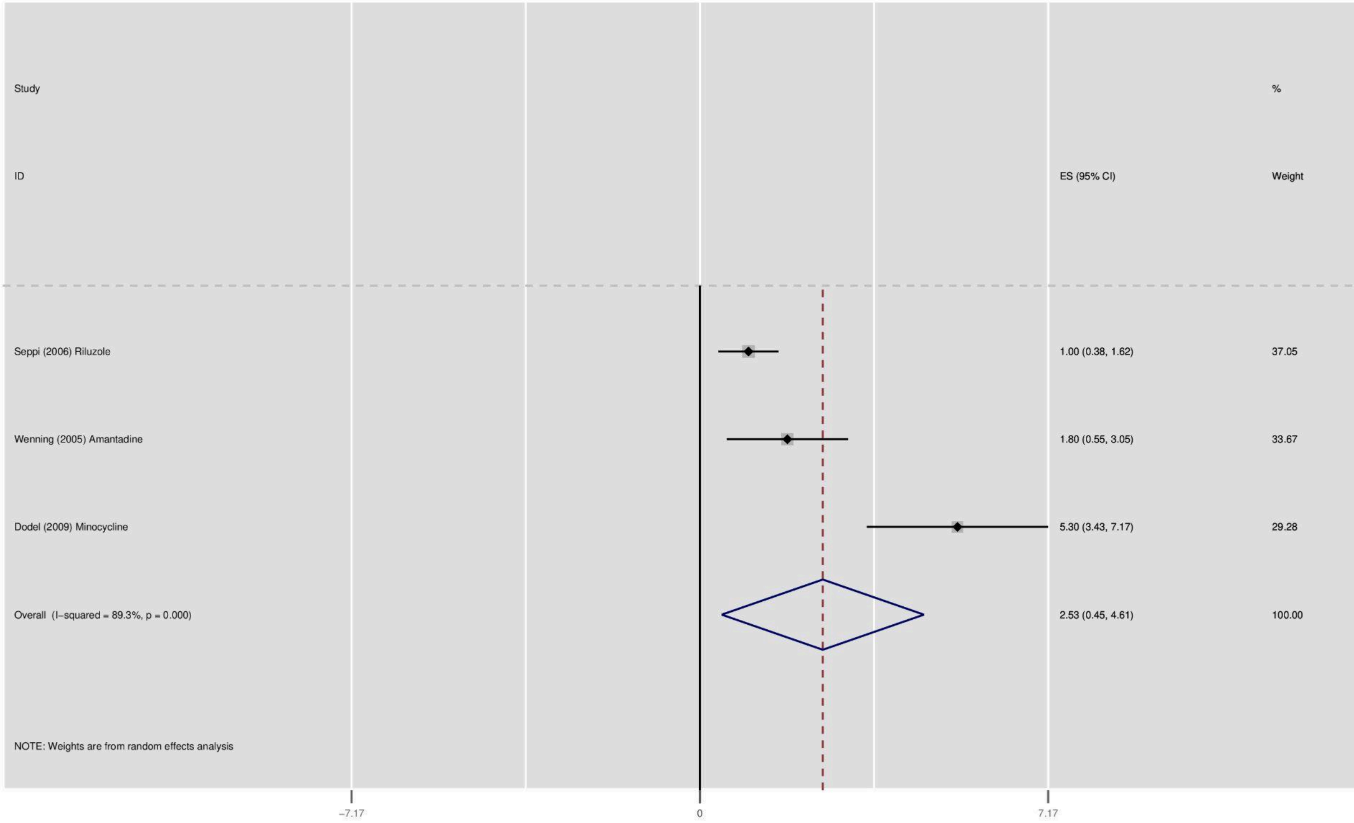
Study
ID
ES (95% CI)
%
Weight
Seppi (2006) Riluzole
1.00 (0.38, 1.62)
37.05
Wenning (2005) Amantadine
1.80 (0.55, 3.05)
33.67
Dodel (2009) Minocycline
5.30 (3.43, 7.17)
29.28
Overall (I−squared = 89.3%, p = 0.000)
2.53 (0.45, 4.61)
100.00
NOTE: Weights are from random effects analysis
−7.17
0
7.17

### Safety Outcomes

#### Subgroup analysis for Primary and Secondary Safety Outcomes

| | Nocebo Response Proportion of Adverse Effects | | | Nocebo Response Serious Adverse Effects Proportion | | | Nocebo Response Withdrawals proportion | | |
|---|---|---|---|---|---|---|---|---|---|
| | Percentage (95% | p value | I2 (%) | Percentage (95% | p value | I2 (%) | Percentage (95% | p value | I2 (%) |
| **Global Results** | | | | | | | | | |
| | 63.88 (41.15 to 8 | p<0.001 | 93.03% | 13.80 (5.77 to 24 | p<0.001 | 74.83% | 13.32 (7.54 to 20 | p<0.001 | 53.51% |
| **Result Consistency** | | | | | | | | | |
| **Publication Status** | | | | | | | | | |
| Published | 92.96 (38.37 to 8 | p<0.001 | 93.61% | 14.89 (6.11 to 26 | p<0.001 | 75.94% | 12.75 (6.74 to 19 | p<0.001 | 56.33% |
| Unpublished | 73.68 (51.21 to 8 | p<0.001 | n/a | 5.26 (0.94 to 24.( | p=0.155 | n/a | 21.05 (8.51 to 43 | p<0.001 | n/a |
| Difference | -12.27 (-97.50 to | p=0.766 | 99.60% | 10.49 (-26.50 to | p=0.787 | 89.25% | -9.50 (-40.51 to 2 | p=0.689 | 79.76% |
| **Data Imputation** | | | | | | | | | |
| Imputation | 70.70 (17.63 to 1 | p=0.001 | 0.00% | n/a | n/a | n/a | n/a | n/a | n/a |
| No Imputation | 61.81 (35.82 to 8 | p<0.001 | 94.06% | n/a | n/a | n/a | n/a | n/a | n/a |
| Difference | 3.33 (-51.52 to 5 | p=0.918 | 99.48% | n/a | n/a | n/a | n/a | n/a | n/a |
| **Study Design** | | | | | | | | | |
| **Follow-Up (weeks)** | | | | | | | | | |
| Coef | 1.40 (0.83 to 1.9 | p<0.001 | 86.46% | 0.51 (0.37 to 0.6 | p<0.001 | 0.00% | 0.27 (0.01 to 0.5 | p=0.050 | 70.32% |
| **Cross-over vs Parallel** | | | | | | | | | |
| Cross-Over | 47.73 (26.11 to 6 | p<0.001 | 0.00% | n/a | n/a | n/a | 1.65 (0.00 to 15. | p=0.506 | 8.30% |
| Parallel | (41.47 to 89.10) | p<0.001 | 94.61% | n/a | n/a | n/a | 15.23 (8.97 to 22 | p<0.001 | 56.81% |
| Difference | -16.88 (-73.07 to | p=0.520 | 99.61% | n/a | n/a | n/a | -11.91 (-28.06 to | p=0.232 | 79.91% |
| **Intervention** | | | | | | | | | |
| Pharmacological | 78.60 (64.72 to 9 | p<0.001 | 81.12% | 19.36 (11.23 to 2 | p<0.001 | 60.33% | 17.77 (12.72 to 2 | p<0.001 | 17.36% |
| Non-Pharmacological | 00 (0.00 to 4.90) | p=1.000 | 0.00% | 00 (0.00 to 4.90) | p=1.000 | 0.00% | 0.00 (0.00 to 4.0 | p=1.000 | 0.00% |
| Difference | 77.63 (47.53 to 1 | p=0.015 | 87.60% | 18.17 (-3.76 to 4 | p=0.140 | 85.64% | 15.54 (2.80 to 28 | p=0.033 | 73.19% |
| **International** | | | | | | | | | |
| International | 78.10 (69.69 to 8 | p<0.001 | 61.23% | 21.74 (3.37 to 48 | p=0.004 | 83.02% | 23.53 (13.25 to 3 | p<0.001 | n/a |
| Non-international | (30.36 to 85.78) | p<0.001 | 93.87% | 10.39 (2.82 to 20 | p<0.001 | 66.02% | 9.941 (3.82 to 17 | p<0.001 | 51.16% |
| Difference | 23.62 (-36.43 to | p=0.363 | 99.59% | 9.87 (-14.34 to 3 | p=0.424 | 88.85% | 15.82 (0.67 to 30 | p=0.018 | 67.46% |
| **Multi-center** | | | | | | | | | |
| Multi-centric | 78.04 (63.19 to 9 | p<0.001 | 81.84% | 19.36 (11.23 to 2 | p<0.001 | 60.33% | 17.79 (11.56 to 2 | p<0.001 | 42.01% |
| Single-Centric | 4 (0.00 to 92.98) | p<0.001 | 95.45% | 00 (0.00 to 4.90) | p=1.000 | 0.00% | 6.73 (0.00 to 19. | p=0.043 | 52.89% |
| Difference | 35.47 (-5.05 to 7 | p=0.130 | 99.24% | 18.17 (-3.75 to 4 | p=0.145 | 85.64% | 8.42 (-4.64 to 21 | p=0.149 | 75.88% |
| **Year of Publication** | | | | | | | | | |
| Coef | 0.04 (-2.66 to 2.7 | p=0.949 | 99.55% | -1.84 (-4.86 to 1. | p=0.281 | 88.36% | -0.51 (-1.52 to 0. | p=0.344 | 79.84% |
| **Allocation Ratio** | | | | | | | | | |
| Coef per % change | -0.01 (-0.03 to 0. | p=0.530 | 99.69% | 0.01 (-0.01 to 0.0 | p=0.322 | 88.98% | 0.00 (0.00 to 0.0 | p=0.279 | 83.59% |
| **Population Characteristics** | | | | | | | | | |
| **% males** | | | | | | | | | |
| Coef per % change | 1.07 (-0.39 to 2.5 | p=0.213 | 99.51% | 0..89 (-0.15 to 1. | p=0.113 | 89.21% | -0.08 (-0.53 to 0. | p=0.757 | 80.81% |
| **Mean Age (years)** | | | | | | | | | |
| Coef | 2.52 (-1.35 to 6.3 | p=0.119 | 99.01% | 2.39 (1.00 to 3.7 | p=0.006 | 53.26% | 0.60 (-0.83 to 2.0 | p=0.376 | 77.87% |
| **Mean Motor Scale (units)** | | | | | | | | | |
| Coef | 0.01 (-2.39 to 2.4 | p=0.940 | 91.76% | -0.14 (-1.49 to 1. | p=0.758 | 43.57% | 0.78 (-0.22 to 1.7 | p=0.160 | 69.01% |

### Bibliography


1. Neal R Haddaway, Luke McGuinness. *PRISMA2020: R Package and ShinyApp for Producing PRISMA 2020 Compliant Flow Diagrams*. Zenodo; 2020. doi:10.5281/ZENODO.4287835
2. Glanville JM. How to identify randomized controlled trials in MEDLINE: ten years on. *J Med Libr Assoc*. Published online 2006:7.
3. Lefebvre C, Eisinga A, McDonald S, Paul N. Enhancing access to reports of randomized trials published world-wide – the contribution of EMBASE records to the Cochrane Central Register of Controlled Trials (CENTRAL) in The Cochrane Library. *Emerg Themes Epidemiol*. 2008;5(1):13. doi:10.1186/1742-7622-5-13
4. Wong; S, Wilczynski; N, Haynes R. Developing optimal search strategies for detecting clinically sound treatment studies. *Med Libr Assoc*. J2006(94(1)):41-47.
5. Higgins J, Altman D. Assessing Risk of Bias in Included Studies. In: *Cochrane Handbook for Systematic Reviews of Interventions:*. ; 2008:187-241.
6. Wasserman S, Hedges LV, Olkin I. Statistical Methods for Meta-Analysis. *J Educ Stat*. 1988;13(1):75. doi:10.2307/1164953
7. Durlak JA. How to Select, Calculate, and Interpret Effect Sizes. *J Pediatr Psychol*. 2009;34(9):917-928. doi:10.1093/jpepsy/jsp004
8. Follmann D, Elliott P, Suh I, Cutler J. Variance imputation for overviews of clinical trials with continuous response. *J Clin Epidemiol*. 1992;45(7):769-773. doi:10.1016/0895-4356(92)90054-Q
9. Abrams KR, Gillies CL, Lambert PC. Meta-analysis of heterogeneously reported trials assessing change from baseline. *Stat Med*. 2005;24(24):3823-3844. doi:10.1002/sim.2423
10. Hozo SP, Djulbegovic B, Hozo I. Estimating the mean and variance from the median, range, and the size of a sample. *BMC Med Res Methodol*. 2005;5(1):13. doi:10.1186/1471-2288-5-13
11. Neal R Haddaway, Luke McGuinness. *PRISMA2020: R Package and ShinyApp for Producing PRISMA 2020 Compliant Flow Diagrams*. Zenodo; 2020. doi:10.5281/ZENODO.4287835
12. Bensimon G, Ludolph A, Agid Y, Vidailhet M, Payan C, Leigh PN. Riluzole treatment, survival and diagnostic criteria in Parkinson plus disorders: The NNIPPS Study. *Brain*. 2009;132(1):156-171. doi:10.1093/brain/awn291
13. Bonnet AM, Esteguy M, Tell G, Schechter PJ, Hardenberg J, Agid Y. A Controlled Study of Oral Vigabatrin (ɣ-Vinyl GABA) in Patients with Cerebellar Ataxia. *Can J Neurol Sci J Can Sci Neurol*. 1986;13(4):331-333. doi:10.1017/S0317167100036672
14. Bordet R, Benhadjali J, Destée A, Belabbas A, Libersa C. Octreotide Effects on Orthostatic Hypotension in Patients with Multiple System Atrophy: A Controlled Study of Acute Administration. *Clin Neuropharmacol*. 1995;18(1):83-89. doi:10.1097/00002826-199502000-00012
15. Chou Y hui, You H, Wang H, et al. Effect of Repetitive Transcranial Magnetic Stimulation on fMRI Resting-State Connectivity in Multiple System Atrophy. *Brain Connect*. 2015;5(7):451-459. doi:10.1089/brain.2014.0325
16. Dodel R, Spottke A, Gerhard A, et al. Minocycline 1-year therapy in multiple-system-atrophy: Effect on clinical symptoms and [ $^{11}$ C] *(R)* -PK11195 PET (MEMSA-trial): *Efficacy of Minocycline for Patients with MSA-P. Mov Disord*. 2010;25(1):97-107. doi:10.1002/mds.22732
17. França C, de Andrade DC, Silva V, et al. Effects of cerebellar transcranial magnetic stimulation on ataxias: A randomized trial. *Parkinsonism Relat Disord*. 2020;80:1-6. doi:10.1016/j.parkreldis.2020.09.001
18. Friess E, Kuempfel T, Modell S, et al. Paroxetine treatment improves motor symptoms in patients with multiple system atrophy. *Parkinsonism Relat Disord*. 2006;12(7):432-437. doi:10.1016/j.parkreldis.2006.04.002

19. Holmberg B, Johansson JO, Poewe W, et al. Safety and tolerability of growth hormone therapy in multiple system atrophy: A double-blind, placebo-controlled study. *Mov Disord*. 2007;22(8):1138-1144. doi:10.1002/mds.21501
20. Kaufmann H, Freeman R, Biaggioni I, et al. Droxidopa for neurogenic orthostatic hypotension: A randomized, placebo-controlled, phase 3 trial. *Neurology*. 2014;83(4):328-335. doi:10.1212/WNL.0000000000000615
21. Lee PH, Lee JE, Kim HS, et al. A randomized trial of mesenchymal stem cells in multiple system atrophy. *Ann Neurol*. 2012;72(1):32-40. doi:10.1002/ana.23612
22. Jung Lee J, Han Yoon J, Jin Kim S, et al. Inosine 5'-Monophosphate to Raise Serum Uric Acid Level in Multiple System Atrophy (IMPROVE-MSA study). *Clin Pharmacol Ther*. Published online November 30, 2020:cpt.2082. doi:10.1002/cpt.2082
23. Levin J, Maaß S, Schuberth M, et al. Safety and efficacy of epigallocatechin gallate in multiple system atrophy (PROMESA): a randomised, double-blind, placebo-controlled trial. *Lancet Neurol*. 2019;18(8):724-735. doi:10.1016/S1474-4422(19)30141-3
24. Low PA, Robertson D, Gilman S, et al. Efficacy and safety of rifampicin for multiple system atrophy: a randomised, double-blind, placebo-controlled trial. *Lancet Neurol*. 2014;13(3):268-275. doi:10.1016/S1474-4422(13)70301-6
25. Meissner WG, Traon AP, Foubert-Samier A, et al. A Phase 1 Randomized Trial of Specific Active A-SYNUCLEIN Immunotherapies PD01A and PD03A in Multiple System Atrophy. *Mov Disord*. 2020;35(11):1957-1965. doi:10.1002/mds.28218
26. Saccà F, Marsili A, Quarantelli M, et al. A randomized clinical trial of lithium in multiple system atrophy. *J Neurol*. 2013;260(2):458-461. doi:10.1007/s00415-012-6655-7
27. Seppi K, Peralta C, Diem-Zangerl A, et al. Placebo-controlled trial of riluzole in multiple system atrophy. *Eur J Neurol*. 2006;13(10):1146-1148. doi:10.1111/j.1468-1331.2006.01452.x
28. Song P, Li S, Wang S, Wei H, Lin H, Wang Y. Repetitive transcranial magnetic stimulation of the cerebellum improves ataxia and cerebello-fronto plasticity in multiple system atrophy: a randomized, double-blind, sham-controlled and TMS-EEG study. *Aging*. 2020;12(20):20611-20622. doi:10.18632/aging.103946
29. Wenning GK. Placebo-Controlled Trial of Amantadine in Multiple-System Atrophy: *Clin Neuropharmacol*. 2005;28(5):225-227. doi:10.1097/01.wnf.0000183240.47960.f0
30. Wessel K, Hermsdorfer J, Deger K, et al. Double-blind Crossover Study With Levorotatory Form of Hydroxytryptophan in Patients With Degenerative Cerebellar Diseases. *Arch Neurol*. 1995;52(5):451-455. doi:10.1001/archneur.1995.00540290037015
31. Poewe W, Seppi K, Fitzer-Attas CJ, et al. Efficacy of rasagiline in patients with the parkinsonian variant of multiple system atrophy: a randomised, placebo-controlled trial. *Lancet Neurol*. 2015;14(2):145-152. doi:10.1016/S1474-4422(14)70288-1
32. AstraZeneca. *A 12-Week, Multicenter, Randomized, Parallel-Group Study to Assess the Safety, Tolerability, Pharmacokinetics, Biomarker Effects, Efficacy, and Effect on Microglia Activation, as Measured by Positron Emission Tomography, of AZD3241 in Subjects With Multiple System Atrophy*. clinicaltrials.gov; 2017. Accessed February 11, 2021. https://clinicaltrials.gov/ct2/show/NCT02388295
33. Song P, Li S, Wang S, Wei H, Lin H, Wang Y. Repetitive transcranial magnetic stimulation of the cerebellum improves ataxia and cerebello-fronto plasticity in multiple system atrophy: a randomized, double-blind, sham-controlled and TMS-EEG study. *Aging*. 2020;12(20):20611-20622. doi:10.18632/aging.103946
34. McGuinness LA, Higgins JPT. Risk-of-bias VISualization (robvis): An R package and Shiny web app for visualizing risk-of-bias assessments. *Res Synth Methods*. 2021;12(1):55-61. doi:10.1002/jrsm.1411
35. Armstrong E, Mathias CJ. The effects of the somatostatin analogue, octreotide, on postural hypotension, before and after food ingestion, in primary autonomic failure. *Clin Auton Res*. 1991;1(2):135-140. doi:10.1007/BF01826210
36. Aschoff JC, Kailer NA, Walter K. [Physostigmine in treatment of cerebellar ataxia]. *Nervenarzt*. 1996;67(4):311-318.

37. Bier JC, Dethy S, Hildebrand J, et al. Effects of the oral form of ondansetron on cerebellar dysfunction. *J Neurol*. 2003;250(6):693-697. doi:10.1007/s00415-003-1061-9
38. Botez MI, Botez-Marquard T, Elie R, Pedraza OL, Goyette K, Lalonde R. Amantadine hydrochloride treatment in heredodegenerative ataxias: a double blind study. *J Neurol Neurosurg Psychiatry*. 1996;61(3):259-264. doi:10.1136/jnnp.61.3.259
39. Chaudhuri KR, Maule S, Thomaides T, Pavitt D, Mathias CJ. Alcohol ingestion lowers supine blood pressure, causes splanchnic vasodilatation and worsens postural hypotension in primary autonomic failure. *J Neurol*. 1993;241(3):145-152. doi:10.1007/BF00868341
40. Chou Y hui, Hickey PT, Sundman M, Song AW, Chen N kuei. Effects of repetitive transcranial magnetic stimulation on motor symptoms in Parkinson disease: a systematic review and meta-analysis. *JAMA Neurol*. 2015;72(4):432-440. doi:10.1001/jamaneurol.2014.4380
41. Fouad-Tarazi FM, Okabe M, Goren H. Alpha sympathomimetic treatment of autonomic insufficiency with orthostatic hypotension. *Am J Med*. 1995;99(6):604-610. doi:10.1016/S0002-9343(99)80246-0
42. Freeman R, Young J, Landsberg L, Lipsitz L. The treatment of postprandial hypotension in autonomic failure with. :8.
43. Freeman R, Landsberg L, Young J. The treatment of neurogenic orthostatic hypotension with 3,4-DL-threo-dihydroxyphenylserine: A randomized, placebo-controlled, crossover trial. *Neurology*. 1999;53(9):2151-2151. doi:10.1212/WNL.53.9.2151
44. Hussain IF. Treatment of erectile dysfunction with sildenafil citrate (Viagra) in parkinsonism due to Parkinson's disease or multiple system atrophy with observations on orthostatic hypotension. *J Neurol Neurosurg Psychiatry*. 2001;71(3):371-374. doi:10.1136/jnnp.71.3.371
45. Isaacson S, Shill HA, Vernino S, Ziemann A, Rowse GJ. Safety and Durability of Effect with Long-Term, Open-Label Droxidopa Treatment in Patients with Symptomatic Neurogenic Orthostatic Hypotension (NOH303). *J Park Dis*. 2016;6(4):751-759. doi:10.3233/JPD-160860
46. Jordan J, Lipp A, Tank J, et al. Catechol-O-Methyltransferase and Blood Pressure in Humans. *Circulation*. 2002;106(4):460-465. doi:10.1161/01.CIR.0000022844.50161.3B
47. Jordan J, Shannon JR, Biaggioni I, Norman R, Black BK, Robertson D. Contrasting actions of pressor agents in severe autonomic failure. *Am J Med*. 1998;105(2):116-124. doi:10.1016/S0002-9343(98)00193-4
48. Kaufmann H, Saadia D, Voustianiouk A, et al. Norepinephrine Precursor Therapy in Neurogenic Orthostatic Hypotension. *Circulation*. 2003;108(6):724-728. doi:10.1161/01.CIR.0000083721.49847.D7
49. Lee P, Kim J, Bang O, Ahn Y, Joo I, Huh K. Autologous Mesenchymal Stem Cell Therapy Delays the Progression of Neurological Deficits in Patients With Multiple System Atrophy. *Clin Pharmacol Ther*. 2008;83(5):723-730. doi:10.1038/sj.clpt.6100386
50. Lipp A, Tank J, Stoffels M, Arnold G, Luft FC, Jordan J. Riluzole and blood pressurein multiple system atrophy. *Clin Auton Res*. 2003;13(4):271-274. doi:10.1007/s10286-003-0100-z
51. Mancini F, Zangaglia R, Cristina S, et al. Double-blind, placebo-controlled study to evaluate the efficacy and safety of botulinum toxin type A in the treatment of drooling in parkinsonism. *Mov Disord*. 2003;18(6):685-688. doi:10.1002/mds.10420
52. Abstracts of the 6th international multiple system atrophy congress. *Clin Auton Res*. 2018;28(1):137-160. doi:10.1007/s10286-018-0501-7
53. Park HJ, Oh SH, Kim HN, Jung YJ, Lee PH. Mesenchymal stem cells enhance α-synuclein clearance via M2 microglia polarization in experimental and human parkinsonian disorder. *Acta Neuropathol (Berl)*. 2016;132(5):685-701. doi:10.1007/s00401-016-1605-6
54. Pollak P, Mallaret M, Gaio JM, Hommel M, Perret J. Blood pressure effects of

apomorphine and domperidone in parkinsonism. *Adv Neurol*. 1987;45:263-266.
55. Singer W, Cheshire W, Fealey R, et al. Randomized Trial of Rifampicin in MSA: Effect on Autonomic Function (S16.006). *Neurology*. 2014;82(10 Supplement):S16.006.
56. Singer W, Sandroni P, Opfer-Gehrking TL, et al. Pyridostigmine Treatment Trial in Neurogenic Orthostatic Hypotension. *Arch Neurol*. 2006;63(4):513. doi:10.1001/archneur.63.4.noc50340
57. Sunwoo MK, Yun HJ, Song SK, et al. Mesenchymal stem cells can modulate longitudinal changes in cortical thickness and its related cognitive decline in patients with multiple system atrophy. *Front Aging Neurosci*. 2014;6. doi:10.3389/fnagi.2014.00118
58. Wang H, Li L, Wu T, et al. Increased cerebellar activation after repetitive transcranial magnetic stimulation over the primary motor cortex in patients with multiple system atrophy. *Ann Transl Med*. 2016;4(6):103-103. doi:10.21037/atm.2016.03.24
